# Subcallosal Cingulate structural connectivity as biomarker for chronic low back pain

**DOI:** 10.1101/2024.09.17.24313765

**Authors:** Evangelia Tsolaki, Wenxin Wei, Michael Ward, Ausaf Bari, Nader Pouratian

**Affiliations:** Department of Neurosurgery, David Geffen School of Medicine at UCLA, Los Angeles, CA, USA; Department of Neurological Surgery, UT Southwestern Medical Center, Dallas, TX, USA

## Abstract

**Background:** Chronic low back pain (CLBP) poses a significant challenge, contributing significantly to the ongoing opioid crisis while also being a leading cause of disability. Although spinal cord stimulation (SCS) stands as the primary FDA-endorsed method for neuromodulatory therapy in CLBP, there remains a subset of patients unresponsive to SCS and others who experience insufficient pain relief over time. In view of the evidence suggesting the critical role of subcallosal cingulate cortex (SCC) connectivity in pain processing, in the current study we investigated the role of the baseline SCC structural as a potential neuroimaging predictive biomarker to identify patients that are likely to benefit from SCS.

**Methods:** Diffusion magnetic resonance imaging scans were acquired in 8 patients with CLBP (mean (SD) age = 70 (10) years; 6 female/2 male, 6 UCLA site, 2 UTSW) before their initial SCS trial. Probabilistic tractography from subject-specific anatomically defined SCC seed regions to the ventral striatum (VS), anterior cingulate cortex (ACC), uncinate fasciculus (UCF) and bilateral medial prefrontal cortex (mPFC) was used to calculate FSL structural probabilistic connectivity in the target network. To explore cross-sectional variations in SCC connectivity related to SCS trial response, we employed a general linear model (GLM) using the SCC probability of connectivity as dependent variable, and the response to the SCS trial as independent variable. We used Pearson correlation to evaluate further the relationships between the critical SCC probability of connectivity and the change in VAS score after the SCS trial. Finally, the role of depression in the treatment outcome was evaluated.

**Results:** Responders to SCS had significantly lower ipsilateral SCC connectivity to mPFC (F1,8 =8.19, p = 0.03) and VS (F1,8 =17.48, p=0.01) on the left hemisphere compared to non-responders. Pearson correlation analysis showed that decreased ipsilateral SCC baseline connectivity to left mPFC (*p=0*.*03*) and VS (*p*=0.01) was correlated with higher improvement in VAS scores. The baseline depression severity did not significantly influence the change in VAS score following the SCS trial. On the other hand, baseline SCC-VS connectivity on the left hemisphere was a significant predictor of change in VAS score (*p=0*.*02*).

**Conclusions:** Our study highlights the important role of SCC connectivity that can serve as a potential biomarker for CLBP stratification and prediction to SCS treatment. These results can reshape our perspective on CLBP management and can serve as early indicator of response to the treatment providing a personalized approach based on the individual’s underlying SCC connectivity.

## 1. Introduction

Chronic low back pain (CLBP) is a major source of disability worldwide^1^ and a significant contributor to the current opioid crisis.^2^ Identifying effective non-opioid therapies for the treatment of CLBP is a top healthcare priority. Each day in the United States more than 90 individuals die from opioid overdose.^3^ In recent years, the hunt for biomarkers in chronic pain has intensified, as interest has grown in personalized/precision medicine techniques, and the global opioid crisis has underscored the need to accelerate the pace of pain research. However, there remain no objective, measurable biomarkers to explain the diversity in response to established treatments for CLBP and to understand subgroups of patients that will respond to existing treatment options. There is a need for biomarkers that can guide patient selection and therapy.

The most common FDA-approved neuromodulatory therapy for CLBP includes spinal cord stimulation (SCS) stimulation.^4,5^ SCS has been shown to provide a good outcome in the treatment of CLBP,^6–9^ but there are still patients who fail to respond to SCS and many patients who do not derive adequate pain relief in the long term.^10,11^ CLBP patients are often grouped as a single entity but differential responses to therapeutic interventions suggest distinct profiles. The inconsistent efficacy of SCS for CLBP may be understood in terms of a conceptualization of chronic pain as a disorder of distinct brain circuits which become involved with the chronification of pain.^12,13^ This view of CLBP as a disorder of brain circuits rather than peripheral or spinal processes, is supported by a growing body of neuroimaging literature. For example, chronification of pain is associated with global changes in brain networks, such as the nucleus accumbens (NAc), medial prefrontal cortex (mPFC), amygdala, and hippocampus.^14–16^ Furthermore, these brain networks are distinct than those associated with acute pain. The conscious experience of pain can be understood in terms of separate somatosensory, cognitive, and affective domains.^17^ While the somatosensory and cognitive domains predominate in acute pain, chronic pain is associated with a relative increase in the affective component.^18–20^ Thus, CLBP itself is a heterogeneous disorder with varying degrees of neuropathic, nociceptive and affective components. This clinical heterogeneity is reflected in the heterogeneity of brain regions involved based on neuroimaging evidence. Specifically, structural, and functional neuroimaging studies have revealed the involvement of several cortical and subcortical brain areas in patients with CLBP. Structural MRI has shown gray and white matter changes in several central structures.^21–24^ Interestingly, when patients with persistent back pain were compared to those who had recovered from subacute back pain, there was increased white matter connections in the mPFC-amygdala-NAc network.^25^ Functional MRI studies have also shown involvement in these brain regions with changes in various interconnected areas.^26–31^ Taken together these studies reveal significant structural and functional changes in patients with CLBP that involve the ACC, mPFC, amygdala and NAc. Interestingly, all these areas are involved in depression. ^32–35^ and importantly, all these areas have direct white matter connectivity to the subcallosal cingulate cortex (SCC) region.^36–39^

The SCC region has been strongly implicated in the pathophysiology of depression^40^ and plays an important role in key features of major depressive disorder, including emotion regulation,^41,42^ reward anticipation,^43^ and anhedonia.^44,45^ The various subregions of the SCC have been shown to be functionally and structurally connected to limbic brain regions involved in emotion as well as pain processing.^46–51^ While the SCC itself is activated during affective pain processing,^52,53^ WM within the SCC region has connectivity with the ACC, mPFC, NAc and amygdala via three major WM tracts: the forceps minor (FM), uncinate fasciculus (UCF), and cingulum bundle (CB).^49^ Taken together, not only does the SCC appear to mediate the emotional responses in chronic pain, but it may also serve as an information hub that interconnects critical nodes in brain circuits mediating the sensory, cognitive, and affective components involved in pain processing. In our prior work we shown that the strength of SCC structural connectivity to mPFC is associated with response to electroconvulsive (ECT) therapy for treatment-resistant depression, supporting its potential role as a clinically significant biomarker of response to neuromodulatory therapies.^54^

Few studies^55–58^ attempted to identify predictive factors for long-term SCS response. However, no previous study attempted to stratify CLBP patients based on the underlying brain connectivity of areas that are critical to pain chronification or correlated this connectivity pattern with the response to the SCS trial. Considering the significant complications (electrode migration, hematoma formation, infection, spinal cord injury, cerebrospinal fluid leak)^59^ that are associated with the SCS trial phase and the duplication of the clinical procedure that SC trial is required, it is critical to investigate potential biomarker for the SCS trial phase that will show the patients who may benefit from it. Research focused on the development of quantitative, objective biomarkers alongside self-reports to aid diagnosis, and predict treatment efficacy is of increasing importance to combat CLBP.^60^ Considering the evidence indicating the significant impact of SCC connectivity on pain processing, we hypothesize that assessing SCC structural connections through specific brain pathways—such as FM, UCF, CB fibers—using probabilistic tractography could serve as an important imaging biomarker for patient stratification and predict their responsiveness to SCS trial.

## 2. Methods and Materials

### 2.1 Participants and Clinical Evaluations

The University of California Los Angeles (UCLA) Institutional Review Board (IRB) approved the study protocol, and all methods were performed in accordance with the UCLA IRB guidelines and regulations. Written informed consents were obtained from all subjects. The same procedure was followed on US Southwestern (UTSW) where the UTSW IRB approved the study. Overall, eight subjects (6 UCLA site, 2 UTSW) (Table 1) were recruited from pain management physicians at UCLA and UTSW. Each patient (≥ 21 years) had been advised by a pain management doctor to undergo a percutaneous SCS trial for persistent back pain, with or without pain extending to the legs. CLBP was defined by persistent low back pain despite prior treatments that continues for 12 weeks or longer. Each subject’s pain level was evaluated using the visual analog scale (VAS) before the SCS and after 15 days after the SCS trial. Response to SCS trial was defined as > 50% reduction on VAS. To evaluate the depression of the patients the Hamilton Depression Rating scale (HAMD) was used. To assess the effect of the SCS trial to the VAS and HAMD scores, the nonparametric Wilcoxon Signed-Rank test was used. Finally, to compare the change in the VAS and HAMD scores between the responders and non-responders a Mann-Whitney U test was conducted.

**Table 1.**
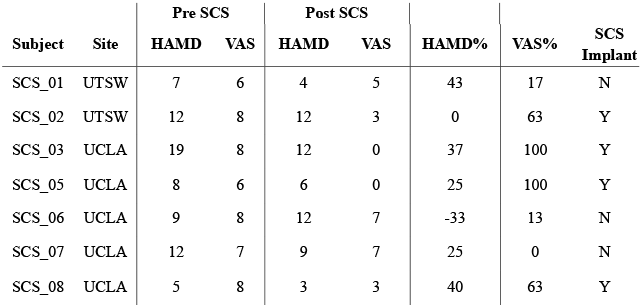
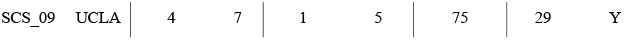
Clinical characteristics.

| Subject | Site | Pre SCS |  | Post SCS |  | HAMD% | VAS% | SCS<br>Implant |
| --- | --- | --- | --- | --- | --- | --- | --- | --- |
|  |  | HAMD | VAS | HAMD | VAS |  |  |  |
| SCS_01 | UTSW | 7 | 6 | 4 | 5 | 43 | 17 | N |
| SCS_02 | UTSW | 12 | 8 | 12 | 3 | 0 | 63 | Y |
| SCS_03 | UCLA | 19 | 8 | 12 | 0 | 37 | 100 | Y |
| SCS_05 | UCLA | 8 | 6 | 6 | 0 | 25 | 100 | Y |
| SCS_06 | UCLA | 9 | 8 | 12 | 7 | -33 | 13 | N |
| SCS_07 | UCLA | 12 | 7 | 9 | 7 | 25 | 0 | N |
| SCS_08 | UCLA | 5 | 8 | 3 | 3 | 40 | 63 | Y |
| SCS_09 | UCLA | 4 | 7 | 1 | 5 | 75 | 29 | Y |

### 2.2 MRI Imaging

Each subject underwent 3T MRI imaging on a Siemens Magentom Prisma scanner prior to SCS trial. The same MRI acquisition protocol was followed by both participating sites (UCLA and UTSW). High resolution T1-weighted anatomical images based on magnetization prepared rapid acquisition gradient echo (MPRAGE) sequences were acquired using the following parameters: TE = 2.24 ms, TR = 2.4 s, matrix = 256 × 256, isotropic 1 mm voxels, and flip angle = 15°. Single shot spin echo planar imaging for Diffusion-weighted MRI data were acquired with the following parameters: 64 directions, FOV=256mm, TR/TE/=4.1ms/7.1ms, multi-slice acceleration=2, b=0/1000 s/mm^2^, Echo Spacing=0.67ms, EPI factor= 12, voxel size 2×2×2mm^3^.We collected this sequence both in the anterior-to-posterior phase encoding direction (AP) and the posterior-to-anterior (PA) direction.

Diffusion-weighted and T1 images were processed with FMRIB’s Software Library (FSL v6.0 www.fmrib.ox.ac.uk/fsl/). For the diffusion data acquisition, reversed phase-encoding directions (AP and PA) resulted in pairs of images with distortions in opposite directions. Prior to tractography, the data were corrected for susceptibility, current-induced distortions, and movement using the FSL topup^61^ and eddy^62^ tools. Subsequently, a multi-fiber diffusion model was implemented in FDT, FMRIB’s Diffusion Toolbox^63^. This model uses Bayesian techniques to estimate a probability distribution function for the principal fiber direction at each voxel, accounting for the possibility of crossing fibers. Three fiber directions were modeled per voxel, using a multiplicative factor of 1 for the prior on the additional modeled fibers and 1,000 iterations before sampling. Diffusion and T1 data were skull stripped using the brain extraction tool ^64^. Finally, T1 images will be segmented^65^ to create a cerebrospinal fluid (CSF) mask to restrict tractography results to brain voxels only and will be linearly registered to diffusion space.

### 2.3 Probabilistic Tractography

Anatomical SCC seed regions were defined as 5×5×5 voxel ROIs centered at x=±6, y=+26, z=-10^66^ in MNI152 (1mm) standard space in the left and right hemisphere as center points. The masks were registered into each individual’s T1 and diffusion imaging space using FLIRT^67^ and FNIRT^68^ for linear and non-linear transformations. Then using the SCC areas as seeds, probabilistic tractography^63^ was performed using FSL FDT tools to delineate the connectivity of the SCC region with the entire brain. The CSF mask will be used as an exclusion mask to avoid the false positive connectivity voxels. Tractography parameters will include 5000 samples, 0.2 curvature threshold, loop check termination, 2000 maximum number of steps, 0.5 mm step length and 0.01 subsidiary fiber fraction threshold. In the resulting whole brain tractography map, the patient-specific white matter pathways (FM, CB, and UCF) were defined in a semi-automated fashion, using methods previously described^69^. For each tract (UCF) or target region (mPFC, ACC, ventral striatum (VS)), larger regions of interest were predefined in a specific range of coordinates as we reported in a previous study.^50^ Then, we evaluated, the regions where the whole brain SCC tract map intersected with these extended areas of interest in order to define subject-specific projection targets. For each anatomically defined target, the coordinates of the voxel with maximum intensity value in the target area was used as a center point to create each patient-specific ROIs. Then, probabilistic tractography was performed to estimate the ipsilateral connectivity from the left and right SCC to subject-specific VS, UCF, and ACC ROIs in and to the left and right mPFC termination regions separately. Each ROI was defined as a waypoint mask in order to discard tracts that do not pass through the target, a termination mask in order to terminate the pathways as soon as they enter the termination mask, and a classification mask in order to quantify connectivity values between the seed and target mask.^70^ The number of the streamlines connecting the SCC with each ROI was estimated and the probability of connectivity of the SCC with each target for VS and ACC, UCF and mPFC in both hemispheres was calculated as the percentage of the streamlines that successfully reach the ROI mask. To compensate for the distance-dependent effect, all the calculated probabilities of connectivity were corrected by the average distance of the seed to the corresponding target area. Finally, each SCC to target probabilistic tract was registered to MNI152 space and the average tract per group (responders, non-responders) was calculated for each target.

### 2.4 Statistical Analysis

To explore cross-sectional variations in SCC connectivity related to SCS trial response (including responders and non-responders), we employed a general linear model (GLM) using the SCC probability of connectivity as dependent variable the response to the SCS trial as independent variable. Although categorizing responses can improve statistical power and aid in clinical decision-making ^71^, setting a response threshold is inherently subjective. Analyzing both dichotomized and continuous responses offers a more comprehensive representation of the relationship between variables. We used Pearson correlation to evaluate further the relationships between SCC probability of connectivity that was found to have significant group differences and the change in VAS score after the SCS trial. Finally, to evaluate the role of depression in the treatment outcome, a multiple regression was run using the critical SCC probability of connectivity and the HAMD baseline score as independent variables and the percentage of improvement in VAS score as dependent variable. The statistical analysis was performed using IBM SPSS (version 29.0, IBM, Armonk, NY).

## 3. Results

### 3.1. Demographics

Demographics are reported to Table 1. No difference in age (*F*_*1*,8_ = 2.33, *p* = 0.18) and gender (χ^2^ =0.53, *p* = 0.46) was observed between the responders (age=72.75 ± 3.30, female=3, male=1) and non-responders (age=63.00 ± 4.12, female=2, male=2). The Wilcoxon Signed-Rank indicated a significant decrease (Z=-2.37, p=0.02) in VAS scores for the whole group of patients between baseline (7.25 ± 0.89) and after SCS trial (3.75 ± 2.76). No significant changes were observed in HAMD scores (Z=-1.64, p=0.10). A Mann-Whitney U test was conducted to compare the change in clinical scores between the responders and non-responders to SCS trial. The results showed that the change in VAS scores were significant higher (Z=-2.337, p=0.29) in responders (81.25 % ± 21.65) compared to non-responders (14.44 % ± 11.79). No significant differences were observed in HAMD scores between the two groups (p>0.05).

### 3.2. Differences in SCC connectivity between responders and non-responders

We conducted a univariate GLM to examine the effect of SCC baseline connectivity to response to SCS trial. We observed that responders had significantly lower ipsilateral SCC probability of connectivity to mPFC L (*F*_*1*,8_ =8.19, *p* = 0.03) and to VS (*F*_*1*,8_ =17.48, *p*=0.01) on the left hemisphere compared to non-responders (*mPFC L*: responders:0.94 ± 0.33, non-responders: 2.36 ± 0.94, *VS*: responders:1.12 ± 0.41, non-responders: 2.29 ± 0.41). (Figure 1A) Pearson correlation analysis examined the change in VAS score after SCS trial and SCC baseline connectivity and showed that decreased ipsilateral SCC connectivity to mPFC L and VS on the left hemisphere was related with higher improvement in VAS scores (mPFC L: r=-0.74, *p=0*.*03*, VS: r=-0.82, *p=0*.*01*). (Figure 1B) The mean tracts of the critical SCC connectivity (mPFC L, VS) for each group are illustrated in Figure 2.

**Figure 1.**
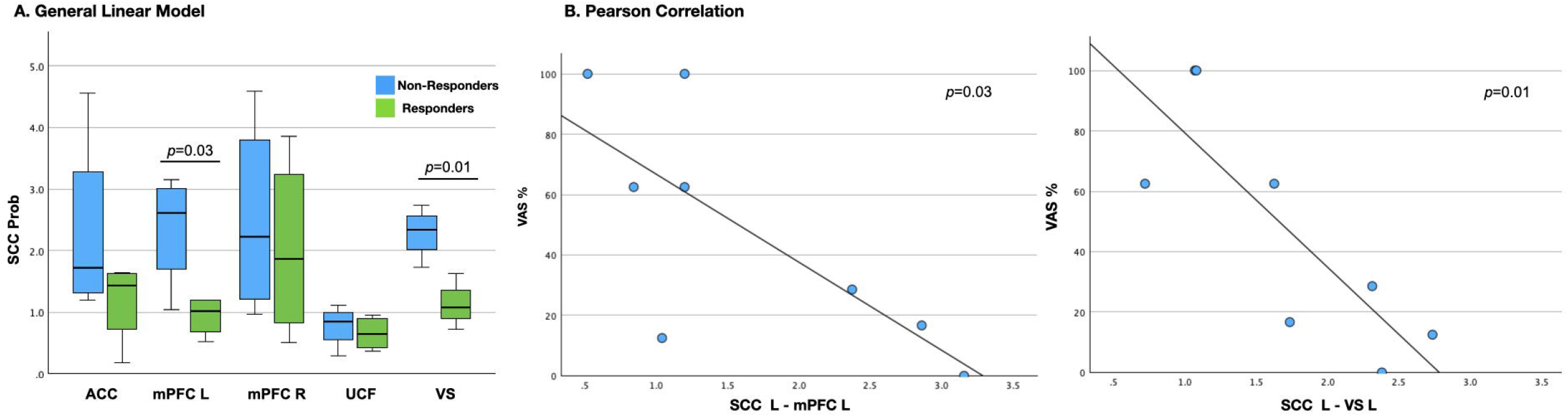
Differentiation between SCS trial responders and non-responders. **(A)** At baseline non-responders to SCS presented significantly higher SCC probability of connectivity (corrected by distance) to ipsilateral mPFC and VS compared to responders on the left hemisphere. **(B)** Pearson correlation analysis showed that the lower ipsilateral SCC probability ofconnectivity tom.PFC and VS at baseline corresponds to higher change in VAS pain score after SCS trial.

**Figure 2.**
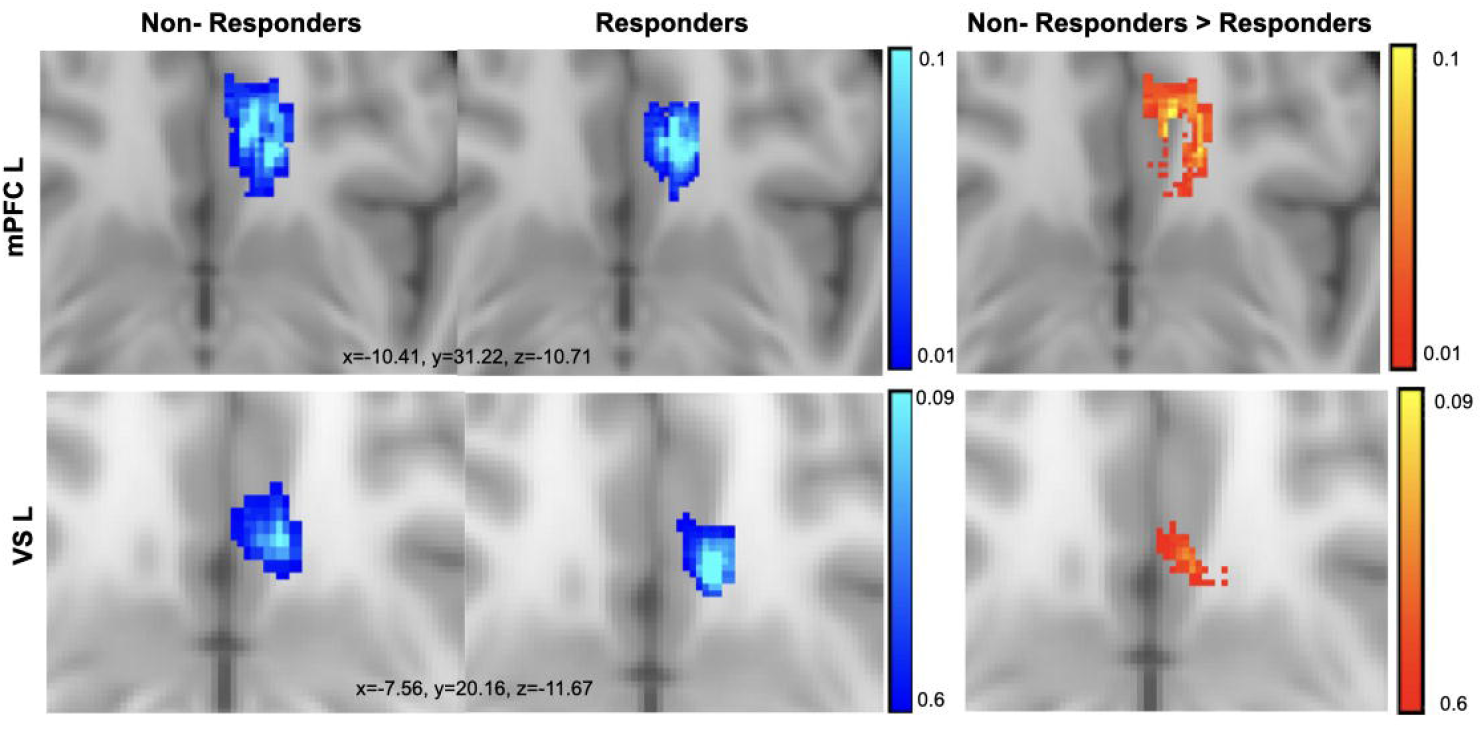
Probabilistic Tractography. The comparison between the average tract across the two groups showed that non-responders presented higher SCC connectivity to mPFC L and VS compared to responders at baseline in line with the statistical analysis. The red tract corresponds to the voxels where the non-responders presented higher SCC structural connectivity compared to responders.

### 3.3. Role of depression in SCS trial outcome

A multiple regression model was performed to evaluate the role of depression in the SCS trial outcome. The relationship between the change in VAS scores with the critical SCC connectivity (mPFC, VS) and the HAMD at baseline was examined. The overall model was statistically significant (F(2, 5) =7.01, p = *0*.*03, R*^*2*^=0.74) only for the SCC-VS connectivity. The baseline HAMD was not found to be a significant predictor of %VAS (b=2.081, t(5)=1.10, *p=0*.*32*), indicating that baseline severity of depression symptoms, as measured by the HAMD scale, did not significantly influence the change in VAS score following the SCS trial. On the other hand, the baseline SCC-VS connectivity on the left hemisphere was a significant predictor of change in VAS score (b=-42.71, t(5)=-25.46, p=0.025). This result indicates that lower baseline ipsilateral SCC-VS connectivity on the left hemisphere associated with higher improvement in VAS scores.

## 4. Discussion

Chronic pain reflects a multidimensional disorder of the brain comprised of sensory, cognitive, and affective components^72^. SCS has been shown to provide a good outcome in the treatment of CLBP,^6–9^ but there are still patients who fail to respond to SCS and many patients who do not derive adequate pain relief in the long term.^10,11^ Considering the significant complications associated with the SCS trial phase and the need to duplicate a clinical procedure, it is essential to identify biomarkers that can predict which patients are likely to benefit from SCS trial. To the best of our knowledge this is the first time that a study attempts to correlate the brain connectivity of areas that are critical to pain chronification connectivity pattern with the response to the SCS trial. Our findings indicate that baseline SCC structural connectivity to mPFC and VS differs between responders and non-responders to SCS trial, suggesting that SCC structural connectivity may serve as a potential neuroimaging biomarker to predict those that are likely to benefit from SCS intervention. Specifically, we found that responders to SCS trial demonstrated significantly lower ipsilateral SCC connectivity to mPFC and VS on the left hemisphere when compared to non-responders. Evaluating the role of the severity of depression at baseline to the treatment outcome, baseline HAMD did not significantly influence the change in VAS following the SCS trial, suggesting that the effectiveness of SCS trial was not dependent on how depressed the patient was at baseline.

The mechanism of action of SCS is poorly understood and clinical outcome is dependent on factors beyond the technical aspects of implantation. Psychological factors have been suggested to be considered when investigating SCS efficacy.^73^ In particular, depression is known to be associated with chronic pain and can influence the outcome of pain therapy.^74^ Frequent comorbidity of depression and chronic pain has inspired the formulation of a hypothesis regarding a shared neurobiological mechanism of both conditions. The bulk of evidence suggests that the pathophysiology of chronic pain and depression is closely coupled with the abnormal function of the brain networks involved in the regulation of both emotions and pain, and other mechanisms involved in these processes such as insufficiency of descending serotonin and noradrenaline pathways and abnormal activation of proinflammatory cytokines and substance P.^75,76^ Even though depression is considered to be the strongest psychological predictor of an unsuccessful SCS,^77^ empirical studies cannot easily identify who will succeed or fail with SCS treatment. Given the high comorbidity, excluding all patients with depression may be overcompensate and withhold treatment from those who may otherwise benefit. Consistent with this, our results demonstrated that baseline depression severity did not significantly affect the change in VAS following the SCS trial, indicating that the effectiveness of the SCS trial was not dependent on the patient’s level of depression at baseline. Considering the significant complications associated with the SCS trial phase, there is a need to understand the factors that contribute to the efficacy of SCS trial considering the CLBP as brain disorder.

The field of pain medicine relies on accurate diagnosis of specific pain conditions. Moreover, it is increasingly recognized that many chronic pain conditions consist of subtypes with distinctly different prognoses and responses to various therapies.^78^ Brain connectivity patterns have been shown to predict the transition from acute to chronic pain.^79–81^ The current findings suggest the potential use of a neuroimaging biomarker that could be used to stratify patients with CLBP. This is particularly important as it could facilitate a more individualized approach to pain treatment. The study’s results may offer deeper insights into the underlying pathology of CLBP, serve as an early indicator of comorbidities that are not yet clinically detectable, and help clinicians determine whether SCS is the optimal treatment based on each patient’s specific brain connectivity pattern. Currently, pain assessment in clinical settings primarily relies on rating scales and symptom-based questionnaires^82–87^. However, these subjective measures are influenced by contextual factors and offer only moderate reliability, even with extensive training programs aimed at improving the accuracy of self-reported pain^88^. This underscores the significant need for universally accepted objective biomarkers for pain. The findings from the current study could provide a potential important complementary tool to self-reports, aiding in the assessment and differentiation of the mechanisms and causes of CLBP. The development of the proposed biomarker could significantly advance the care of CLBP patients and play a pivotal role in precision pain medicine.

Despite the significant findings, several limitations need to be addressed. The primary limitation is the small sample size, which reduces the study’s statistical power and limits the generalizability of the results. A larger sample size would allow for more robust conclusions and improve the reliability of the observed associations. Future research should involve larger, more diverse patient cohorts to confirm and extend these findings. Another limitation is the absence of a harmonization process for the data obtained from different MRI scanners at UCA and UTSW, which may introduce variability and affect the comparability of results across sites. Differences in scanner models, settings, and protocols can influence imaging data, potentially confounding the study’s results. Implementing a comprehensive data harmonization process in future studies would help minimize variability across scanners, enhancing the reliability and reproducibility of the findings Finally, other potential confounding factors, such as patient comorbidities or medication use, were not controlled for in this study, and should be accounted for in future research to ensure the findings are not biased by these variables. Addressing these limitations will be crucial for translating the findings into clinical practice.

## 5. Conclusion

Our study highlights the important role of SCC connectivity that can serve as a potential biomarker for CLBP stratification and prediction to SCS treatment. These results can reshape our perspective on CLBP management and can serve as early indicator of response to the treatment providing a personalized approach based on the individual’s underlying SCC connectivity.

## Data Availability

All data produced in the present study are available upon reasonable request to the author.

